# An efficient approach to nowcasting the time-varying reproduction number

**DOI:** 10.1101/2023.10.30.23297251

**Authors:** Bryan Sumalinab, Oswaldo Gressani, Niel Hens, Christel Faes

**Affiliations:** Interuniversity Institute for Biostatistics and statistical Bioinformatics (I-BioStat), Data Science Institute (DSI), Hasselt University, Hasselt, Belgium; Department of Mathematics and Statistics, College of Science and Mathematics, Mindanao State University - Iligan Institute of Technology, Iligan City, Philippines; Centre for Health Economic Research and Modelling Infectious Diseases (CHERMID), Vaccine & Infectious Disease Institute, Antwerp University, Antwerp, Belgium

**Keywords:** Nowcasting, Reproduction number, Reporting delay, Epidemic

## Abstract

Estimating the instantaneous reproduction number (*ℛ*_*t*_) in near real-time is crucial for monitoring and responding to epidemic outbreaks on a daily basis. However, such estimates often suffer from bias due to reporting delays inherent in surveillance systems. A fast and flexible Bayesian methodology is proposed to overcome this challenge by estimating *ℛ*_*t*_ while taking into account reporting delays. Furthermore, the uncertainty associated with the nowcasting of cases is naturally taken into account to get a valid uncertainty estimation of the nowcasted reproduction number. The proposed methodology is evaluated through a simulation study and applied to COVID-19 incidence data in Belgium.

## 1 Introduction

The instantaneous reproduction number (*ℛ*_*t*_) is one of the key infectious disease parameters that can be used to measure how likely a disease is to spread and can help direct effective management tactics during an epidemic. It is defined as the “average number of infections caused by an infectious individual at a given time *t* “. Several methods have been developed to estimate *ℛ*_*t*_ from epidemiological data. A recent paper by Gostic et al. (2020) offers a comprehensive comparison of some established methodologies, discussing challenges and offering recommendations for accurate estimation of the reproduction number. A key issue highlighted by Gostic et al. (2020) is the potential bias in estimating *ℛ*_*t*_ due to delays, such as those between infection and the confirmation of cases. A simple way to adjust for the delay is by sampling from the delay distribution (assumed to be known) and subtracting the sample from the observed data or by shifting estimates/observations backward by approximately the mean of the delay distribution. However, in practice, these approaches are less accurate. Analogous issues arise when there is a delay in the reporting of cases, leading to right truncation of observed data. This impacts the quality of *ℛ*_*t*_ estimates as the reported (observed/available) cases are relatively low compared to actual cases.

One remedy to address this right truncation problem is by using nowcasting techniques (Höhle and an der Heiden, 2014; Van de Kassteele et al., 2019; McGough et al., 2020; Sumalinab et al., 2022). Their aim is to estimate the actual number of new cases by combining the (predicted) not-yet-reported cases and already reported cases. Subsequently, the estimated/nowcasted cases can be used alongside established *ℛ*_*t*_ estimation methods to nowcast *ℛ*_*t*_ values. However, these procedures ignore the uncertainty in the nowcasted cases when estimating *ℛ*_*t*_. Abbott et al. (2020) account for this uncertainty by approximating the reporting delay distribution through the use of bootstrap techniques. The drawback of such an approach is the added level of complexity in implementation and the extended computational time required. This can pose challenges for timely and daily updates of the epidemic situation. We propose to jointly estimate the time-varying lag in reporting and time series of cases to directly derive the reproduction number (including uncertainty quantification) using a fast and flexible approach in a fully Bayesian framework. The proposed methodology jointly models the delay feature and *ℛ*_*t*_ in a data-driven way, making it simpler and computationally more efficient compared to the approach of Abbott et al. (2020). In particular, we combine the Laplacian-P-splines methodology for nowcasting and estimation of *ℛ*_*t*_ proposed by Sumalinab et al. (2022) and Gressani et al. (2022), respectively. The R code used to implement the simulation study is available on GitHub at https://github.com/bryansumalinab/Rnowcasting.git.

## 2 Methodology

### Nowcasting

Let *y*_*t,d*_ represent the incidence cases at time *t* = 1, 2, …, *T* reported with a delay of *d* = 0, 1, …, *D* days. Following Sumalinab et al. (2022), *y*_*t,d*_ is assumed to follow a negative binomial distribution with mean *μ*_*t,d*_, overdispersion parameter *ϕ* and variance 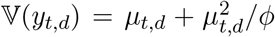. The (log) mean number of cases is modeled using two-dimensional B-splines

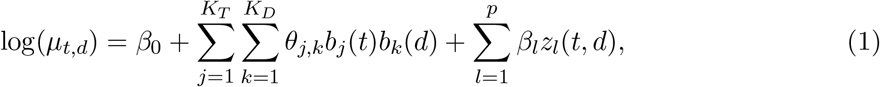

where *β*_0_ is the intercept; *b*_*j*_(*·*) and *b*_*k*_(*·*) are univariate (cubic) B-splines basis functions specified in the time and delay dimensions, respectively and *z*_*l*_(*t, d*) represents day of the week effects with regression coefficients *β*_*l*_.

To control the degree of roughness or smoothness of the fit, we adopt the penalized B-splines (P-splines) methodology proposed by Eilers and Marx (1996). The idea is to specify a substantial number of B-spline basis functions and to counterbalance the implied flexibility by applying a discrete roughness penalty to adjacent B-spline coefficients. This penalty can then be translated into a Bayesian framework by specifying a Gaussian prior distribution for the B-spline coefficients (Lang and Brezger, 2004). Moreover, Gamma priors are assumed for the remaining model hyperparameters. Let ***ξ*** = (***β***^*′*^, ***θ***^*′*^)^*′*^ denote the latent vector with regression coefficients ***β*** = (*β*_0_, *β*_1_, …, *β*_*p*_)^*′*^ and spline parameters 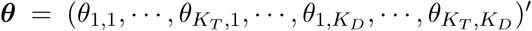. As shown in Sumalinab et al. (2022), the Laplace approximation can be used to approximate the conditional posterior distribution of the latent vector ***ξ*** yielding a Gaussian density with mean 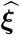 and covariance matrix 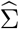. From (1), the posterior mean of the nowcasted cases at time *t* can be derived as

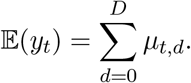

We redirect the reader to the latter reference for full technical details regarding the sampling-free estimation scheme with Laplacian-P-splines

### Estimation of *ℛ*_*t*_

For estimation of the time-varying reproduction number, we follow the work of Gressani et al. (2022) where *ℛ*_*t*_ is expressed as the ratio of the mean incidence divided by the total infectiousness at time *t* (Fraser, 2007) given by

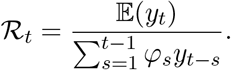

In the above equation, 𝔼(*y*_*t*_) is the mean incidence at time *t* and ***φ*** = (*φ*_1_, …, *φ*_*k*_) is the probability mass function of the serial interval distribution, where *φ*_*s*_ is defined as the probability that the serial interval, i.e. the time elapsed between the onset of symptoms in an infector and the onset of symptoms in the secondary cases generated by that infector, is *s* days. Replacing 𝔼(*y*_*t*_) and *y*_*t−s*_ with the mean nowcasted incidence at time *t* and *t − s*, respectively, the log of *ℛ*_*t*_ is modeled as

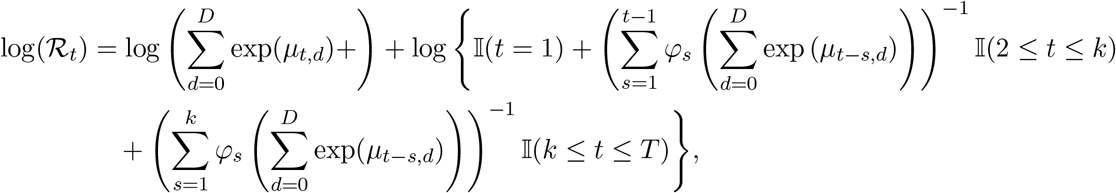

where 𝕀(*·*) is an indicator function such that 𝕀(*A*) equals 1 if *A* is true and 0 otherwise. The estimated time-varying reproduction number 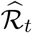 can be derived by substituting *μ*_*t,d*_ by the estimate 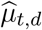 obtained from (1) and the delta method is used to approximate the variance of *ℛ*_*t*_ given by

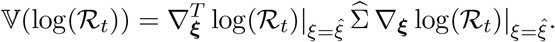

Note that log(*ℛ*_*t*_) is approximately normally distributed with mean 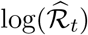 and variance 𝕍(log(*ℛ*_*t*_)). Hence, quantile-based credible intervals for log(*ℛ*_*t*_) can easily be obtained and subsequently, approximate credible intervals for *ℛ*_*t*_ by using the appropriate transformation. A detailed derivation of 𝕍(log(*ℛ*_*t*_)) is given in Appendix A1.

### Simulation study

To assess the performance of our method, a simulation study is conducted and we focus our evaluation on time *T*, that is, the nowcast date. We make certain assumptions on the serial interval distribution and the shape of the true time-varying reproduction number. The latter assumptions enable us to simulate daily case counts according to a negative binomial process. To incorporate reporting delays in the simulation, we assume fixed delay probabilities denoted by *p*_0_, *p*_1_, *p*_2_, …, *p*_*D*_. Subsequently, we generate samples from a multinomial distribution expressed as (*y*_*t*,0_, *y*_*t*,1_, *y*_*t*,2_, …, *y*_*t,D*_) *∼* Multinomial(*y*_*t*_, *p*_0_, *p*_1_, *p*_2_, *· · ·, p*_*D*_). This sample represents the reported number of cases for each (*t, d*) combination. Figure 1 shows the plot of the target reproduction number to be estimated. We choose to focus on several nowcast dates as represented by the dashed vertical lines in Figure 1 with the following characteristics: (i) *T* = 124 - start of the upward trend; (ii) *T* = 140 - increasing phase; (iii) *T* = 150 - local peak; (iv) *T* = 151 - start of the downward trend; (v) *T* = 165 - decreasing phase; and (vi) *T* = 190 - stabilizing phase.

**Figure 1:**
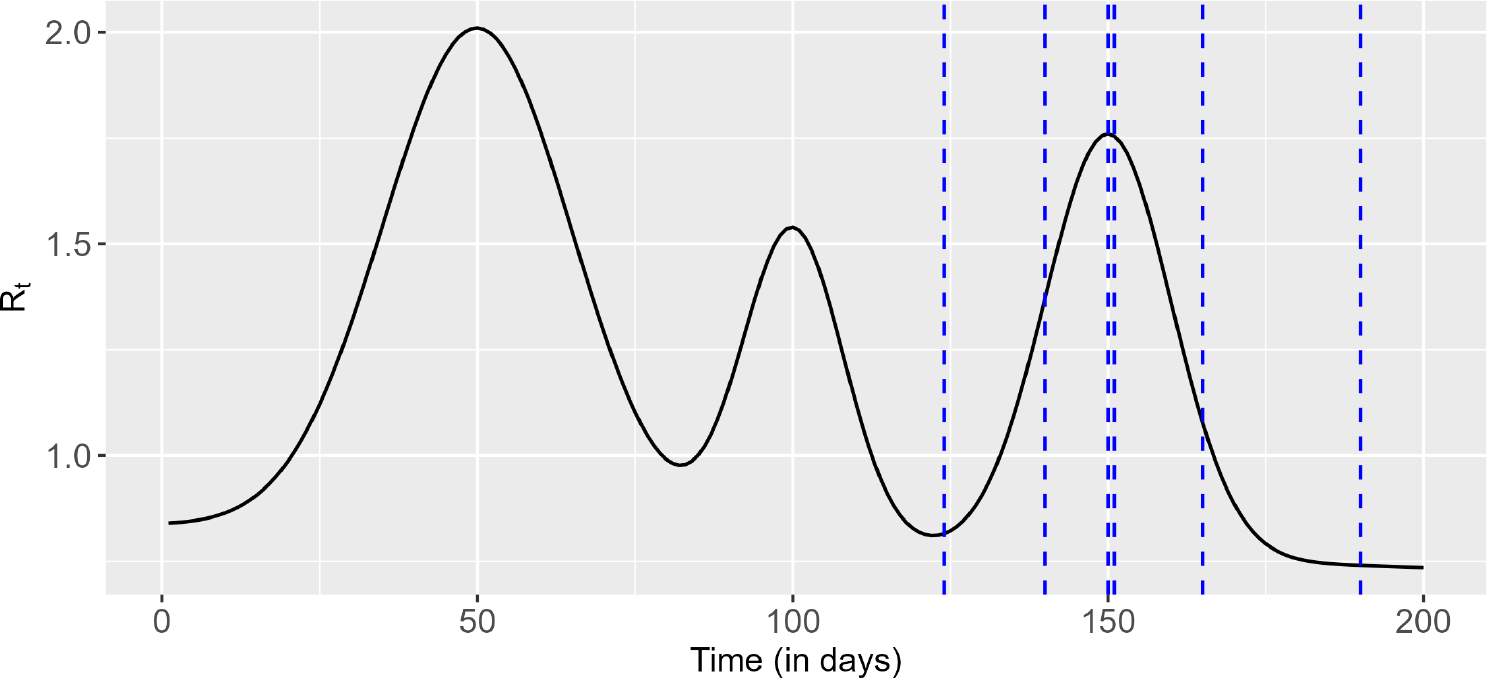
Target time-varying reproduction number with dashed vertical lines corresponding to different nowcast dates of interest.

Several performance measures are considered such as bias, absolute percentage error (APE), credible interval coverage, credible interval width (CI width), and percentage of *ℛ*_*t*_ estimates falling below the target reproduction number 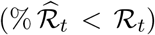. These metrics are computed on the nowcast date, with the median bias, APE, and CI width over 500 simulations being used. Besides assessing the performance of our approach, we also check the performance of the methodology proposed by Gressani et al. (2022) to estimate the instantaneous reproduction number, assuming two different scenarios for the data stream input. A first scenario assumes that the data input solely relies on the reported incidence cases and ignores the nowcasted incidence. A second scenario uses the nowcasted incidence as data input. The former data stream ignores the reporting delays, while the latter takes these delays into account but ignores the uncertainty associated with the delay dimension. In both scenarios, estimation of *ℛ*_*t*_ is carried out using the *estimR()* routine of the EpiLPS package (Gressani, 2021). To summarize, the simulation study allows to compare three models, namely *ℳ*_1_ providing *ℛ*_*t*_ estimates using reported cases only, *ℳ*_2_ providing *ℛ*_*t*_ estimates using nowcasted incidence data and *ℳ*_3_, our proposed model, based on joint modeling of delay and the time-varying reproduction number.

Simulation results are shown in Table 1 assuming no case is reported on the nowcast day. Results solely relying on the reported incidence (*ℳ*_1_) show consistent negative bias and the largest percentage error across all scenarios. This indicates that the target *ℛ*_*t*_ is underestimated when using reported cases only, as expected. Furthermore, it exhibits the highest percentage error, the widest credible interval, and the lowest coverage across all scenarios. On the other hand, when accounting for the reporting delays, both *ℳ*_2_ and *ℳ*_3_ demonstrate similar performance in terms of bias and percentage error, suggesting that point estimates of *ℛ*_*t*_ are relatively close to each other. When *ℛ*_*t*_ is continuously increasing (*T* = 140) or decreasing (*T* = 165), the percentage error is slightly larger (16-18% and 25-27%, respectively) compared to other nowcast dates with errors of less than 10%. Looking at the percentage of *ℛ*_*t*_ estimates falling below the target reproduction number 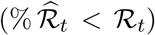, point estimates are mostly below or above the target *ℛ*_*t*_ for increasing and decreasing trends, respectively. The width of the credible interval gets larger when the uncertainty linked to the delays is taken into consideration (*ℳ*_3_), resulting in the highest level of coverage. However, neglecting this uncertainty (*ℳ*_2_) yields a significant undercoverage, suggesting that the constructed credible interval often excludes the target *ℛ*_*t*_. Moreover, the credible interval is widest when *ℛ*_*t*_ is at the local peak (*T* = 150) or in its neighborhood (*T* = 151).

**Table 1:** Performance measures on the nowcast day with no case reported on the nowcast day and delay probabilities (*p*_0_, *p*_1_, *p*_2_, *· · ·, p*_7_) = (0.0, 0.30, 0.25, 0.20, 0.10, 0.10, 0.05, 0.05).

| | Bias | % $\widehat{\mathcal{R}}_t < \mathcal{R}_t$ | APE | 95% CI width | 95% CI Coverage |
| --- | --- | --- | --- | --- | --- |
| **$T = 124$** |  |  |  |  |  |
| $\mathcal{M}_1$ | -0.590 | 100.000 | 72.328 | 0.160 | 2.941 |
| $\mathcal{M}_2$ | 0.025 | 41.176 | 8.873 | 0.231 | 76.471 |
| $\mathcal{M}_3$ | 0.002 | 50.000 | 7.778 | 0.691 | 100.000 |
| **$T = 140$** |  |  |  |  |  |
| $\mathcal{M}_1$ | -0.955 | 100.000 | 69.601 | 0.264 | 2.128 |
| $\mathcal{M}_2$ | -0.212 | 90.213 | 16.390 | 0.286 | 38.723 |
| $\mathcal{M}_3$ | -0.256 | 96.400 | 18.652 | 0.822 | 90.000 |
| **$T = 150$** |  |  |  |  |  |
| $\mathcal{M}_1$ | -1.139 | 100.000 | 64.691 | 0.291 | 0.420 |
| $\mathcal{M}_2$ | -0.061 | 63.866 | 7.272 | 0.250 | 52.101 |
| $\mathcal{M}_3$ | -0.100 | 73.200 | 7.787 | 1.077 | 100.000 |
| **$T = 151$** |  |  |  |  |  |
| $\mathcal{M}_1$ | -1.128 | 100.000 | 64.265 | 0.310 | 0.866 |
| $\mathcal{M}_2$ | -0.018 | 54.113 | 8.252 | 0.236 | 49.351 |
| $\mathcal{M}_3$ | -0.050 | 63.600 | 7.570 | 1.097 | 99.600 |
| **$T = 165$** |  |  |  |  |  |
| $\mathcal{M}_1$ | -0.670 | 100.000 | 62.243 | 0.327 | 1.271 |
| $\mathcal{M}_2$ | 0.280 | 1.695 | 25.975 | 0.135 | 13.983 |
| $\mathcal{M}_3$ | 0.283 | 4.800 | 27.288 | 0.837 | 78.000 |
| **$T = 190$** |  |  |  |  |  |
| $\mathcal{M}_1$ | -0.472 | 100.000 | 63.788 | 0.186 | 2.092 |
| $\mathcal{M}_2$ | 0.010 | 43.515 | 6.724 | 0.170 | 72.385 |
| $\mathcal{M}_3$ | 0.001 | 49.600 | 7.362 | 0.457 | 100.000 |

Additional simulation results are presented in Appendix A2 assuming available data on the nowcast day. Specifically, we assume that 25% and 50% of the cases are reported on the nowcast day. Performance measures are reported in tables 3-4 in Appendix A2. The previously discussed findings in Table 1 remain consistent in the latter simulation scenarios. Furthermore, when examining the results for *ℳ*_3_ in tables 3-4, we observe that the bias and percentage error are quite similar to those in Table 1 except when *ℛ*_*t*_ is at the (local) peak (*T* = 165). There, we observe a decrease in bias and percentage error by 9% and 8%, respectively. This implies that our method performs well in terms of point estimation even when no data is available for the nowcast day. Moreover, the credible interval width is narrower in tables 3-4 and the coverage is closer to the nominal level. This is due to the fact that having more data on the nowcast day results in less uncertainty.

### Real data application

Our methodology is now applied to COVID-19 incidence data in Belgium for the year 2022. The raw data is available on the website of the Sciensano research institute (see https://epistat.sciensano.be/covid/). The data is truncated to a maximum delay of five days and we use the (discrete) serial interval distribution ***φ*** = (0.344, 0.316, 0.168, 0.104, 0.068) from Gressani et al. (2022). Models are fitted with and without day-of-the-week effects and different nowcast dates are compared. Results of these comparisons are available in Appendix A3 and results for the nowcast date of July 31, 2022 are presented in Table 2. The smooth plot of *ℛ*_*t*_ estimates for this nowcast date is presented in Appendix A3. We see in Table 2 that when accounting for reporting delays (*ℳ*_2_ and *ℳ*_3_), both with and without day-of-the-week effects, point estimates are generally closer. Moreover, incorporating day-of-the-week effects for model *ℳ*_3_ leads to a slightly narrower credible interval, which is also observed for the other nowcast dates presented in Appendix A2. This suggests that the inclusion of day-of-the-week effects improves estimation accuracy.

**Table 2:** *ℛ*_*t*_ estimates for the nowcast date on July 31, 2022 using COVID-19 data in Belgium and assessment of day-of-the-week effects. Labels *week* and *no week* represent scenarios with and without day-of-the-week effects, respectively.

| | $\widehat{\mathcal{R}}_t$ | 95% CI lower | 95% CI upper | CI width |
| --- | --- | --- | --- | --- |
| $\mathcal{M}_1$ | 0.780 | 0.634 | 0.960 | 0.326 |
| $\mathcal{M}_2$ - week | 0.899 | 0.747 | 1.081 | 0.335 |
| $\mathcal{M}_2$ - no week | 0.931 | 0.776 | 1.116 | 0.340 |
| $\mathcal{M}_3$ - week | 0.908 | 0.731 | 1.128 | 0.398 |
| $\mathcal{M}_3$ - no week | 0.912 | 0.720 | 1.155 | 0.435 |

## 3 Conclusion

This paper introduces a new methodology for nowcasting the instantaneous reproduction number taking into account reporting delays. The proposed model is fully Bayesian and uses Laplace approximations to posterior distributions as a surrogate to classic Markov chain Monte Carlo techniques to carry out inference. This yields efficient algorithms requiring low computational resources that are particularly well suited for near real-time monitoring of the time-varying reproduction number.

Through the use of Laplacian-P-splines, the methodology presented here shares the same skeleton as the EpiLPS methodology (Gressani et al., 2022), and hence its integration in the latter ecosystem can be done without great difficulty. As such, we plan to integrate nowcasting routines in the EpiLPS package to make them available in a user-friendly environment. It is also worth mentioning that using nowcasted case incidence data as an input in EpiLPS routines (as in *ℳ*_2_) transforms EpiLPS from a retrospective toolbox to a real-time toolbox, at least at the nowcasted time points.

Simulation results show that relying solely on reported cases leads to a significant under-estimation of *ℛ*_*t*_, as well as non-negligible undercoverage. Using the nowcasted incidence without accounting for the time lag uncertainty provides better point estimates of *ℛ*_*t*_ closer to the target value but still with moderate to strong undercoverage. However, directly nowcasting the reproduction number results in better uncertainty quantification as translated by an improved coverage. This can be attributed to wider credible intervals induced by the uncertainty in the reporting delay. Moreover, when data is available on the nowcast date, *ℛ*_*t*_ estimates become more accurate, and the credible intervals have close to nominal value coverage.

## Data Availability

The data is available on the website of the Sciensano Research Institute at https://epistat.sciensano.be/covid/.

### Appendix

#### A1. Derivation for the variance of log(*ℛ*_*t*_)

Note that the log of *ℛ*_*t*_ can be written as:

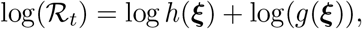

where

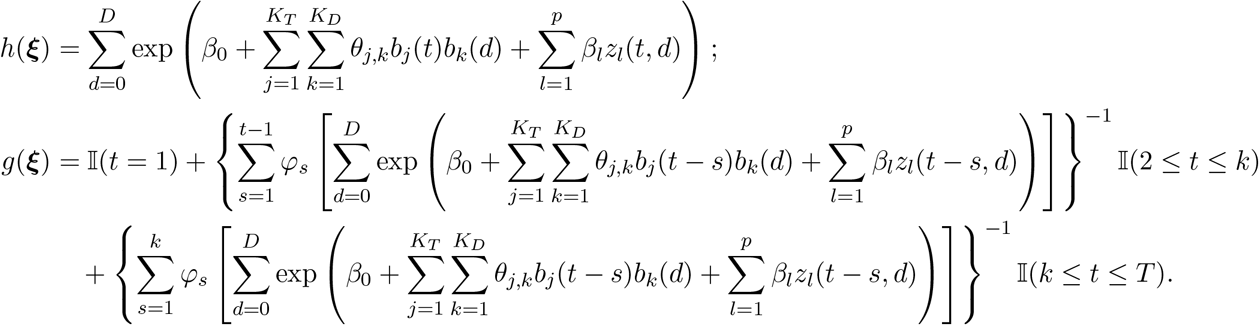

The partial derivatives of *h*(***ξ***) and *g*(***ξ***) with respect to the coefficients *θ*_*j,k*_ and *β*_*l*_ are given by

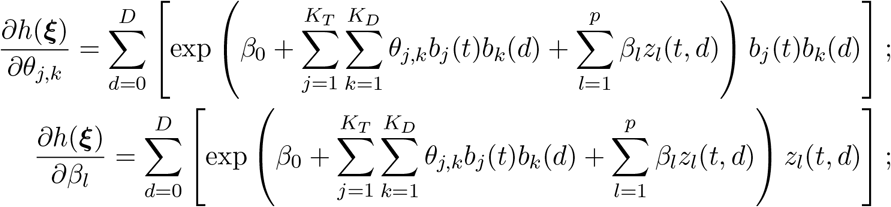

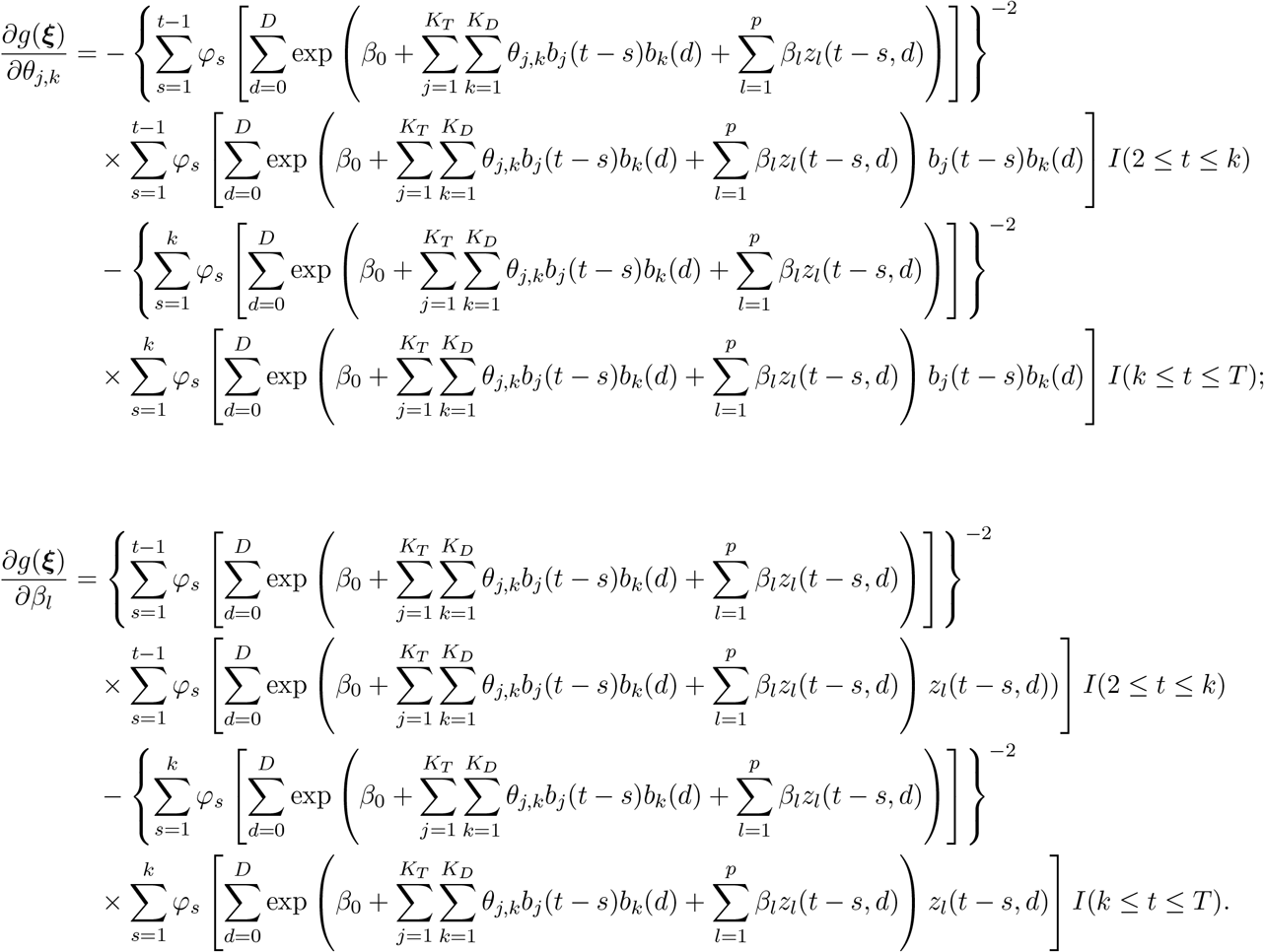

The gradient of log(*ℛ*_*t*_) with respect to ***ξ*** is given by

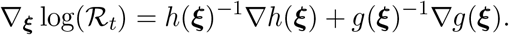

The variance of log(*ℛ*_*t*_) can be approximated as

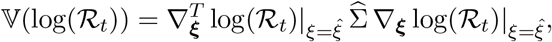

where 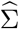 is the estimated covariance matrix of the Laplace approximation for ***ξ***.

#### A2. Additional simulation results

**Table 3:** Performance measures on the nowcast day with 25% of cases reported on the nowcast day and delay probabilities (*p*_0_, *p*_1_, *p*_2_, *· · ·, p*_7_) = (0.25, 0.20, 0.15, 0.15, 0.10, 0.05, 0.05, 0.05).

| | Bias | % $\widehat{\mathcal{R}}_t < \mathcal{R}_t$ | APE | 95% CI width | 95% CI Coverage |
| --- | --- | --- | --- | --- | --- |
| **$T = 124$** |  |  |  |  |  |
| $\mathcal{M}_1$ | -0.439 | 99.583 | 53.845 | 0.172 | 5.000 |
| $\mathcal{M}_2$ | -0.002 | 51.250 | 7.820 | 0.228 | 78.750 |
| $\mathcal{M}_3$ | -0.009 | 55.020 | 9.009 | 0.441 | 95.984 |
| **$T = 140$** |  |  |  |  |  |
| $\mathcal{M}_1$ | -0.723 | 100.000 | 52.728 | 0.261 | 3.347 |
| $\mathcal{M}_2$ | -0.181 | 85.774 | 13.631 | 0.311 | 49.791 |
| $\mathcal{M}_3$ | -0.196 | 90.763 | 14.353 | 0.550 | 76.707 |
| **$T = 150$** |  |  |  |  |  |
| $\mathcal{M}_1$ | -0.828 | 100.000 | 47.048 | 0.216 | 1.702 |
| $\mathcal{M}_2$ | -0.039 | 58.723 | 6.921 | 0.238 | 53.191 |
| $\mathcal{M}_3$ | -0.063 | 63.454 | 7.601 | 0.679 | 93.173 |
| **$T = 151$** |  |  |  |  |  |
| $\mathcal{M}_1$ | -0.831 | 100.000 | 47.376 | 0.205 | 0.000 |
| $\mathcal{M}_2$ | -0.021 | 52.321 | 7.291 | 0.224 | 44.726 |
| $\mathcal{M}_3$ | -0.035 | 57.028 | 7.532 | 0.681 | 94.779 |
| **$T = 165$** |  |  |  |  |  |
| $\mathcal{M}_1$ | -0.505 | 99.569 | 46.877 | 0.126 | 1.293 |
| $\mathcal{M}_2$ | 0.163 | 7.759 | 15.145 | 0.140 | 23.707 |
| $\mathcal{M}_3$ | 0.175 | 10.442 | 17.084 | 0.478 | 64.659 |
| **$T = 190$** |  |  |  |  |  |
| $\mathcal{M}_1$ | -0.352 | 100.000 | 47.500 | 0.130 | 2.146 |
| $\mathcal{M}_2$ | -0.001 | 50.644 | 6.870 | 0.143 | 69.957 |
| $\mathcal{M}_3$ | -0.004 | 51.807 | 8.164 | 0.286 | 93.173 |

**Table 4:** Performance measures on the nowcast day with 50% of cases reported on the nowcast day and delay probabilities (*p*_0_, *p*_1_, *p*_2_, *· · ·, p*_7_) = (0.50, 0.10, 0.10, 0.10, 0.05, 0.05, 0.05, 0.05).

| | Bias | % $\widehat{\mathcal{R}}_t < \mathcal{R}_t$ | APE | 95% CI width | 95% CI Coverage |
| --- | --- | --- | --- | --- | --- |
| **$T = 124$** |  |  |  |  |  |
| $\mathcal{M}_1$ | -0.304 | 99.578 | 37.247 | 0.189 | 8.861 |
| $\mathcal{M}_2$ | 0.008 | 46.835 | 8.213 | 0.224 | 75.105 |
| $\mathcal{M}_3$ | -0.012 | 53.012 | 8.786 | 0.494 | 99.598 |
| **$T = 140$** |  |  |  |  |  |
| $\mathcal{M}_1$ | -0.503 | 100.000 | 36.691 | 0.273 | 5.579 |
| $\mathcal{M}_2$ | -0.166 | 90.129 | 12.276 | 0.299 | 51.073 |
| $\mathcal{M}_3$ | -0.197 | 92.369 | 14.447 | 0.631 | 85.542 |
| **$T = 150$** |  |  |  |  |  |
| $\mathcal{M}_1$ | -0.540 | 100.000 | 30.678 | 0.233 | 3.814 |
| $\mathcal{M}_2$ | -0.039 | 57.627 | 7.007 | 0.244 | 54.661 |
| $\mathcal{M}_3$ | -0.126 | 71.888 | 8.416 | 0.774 | 96.386 |
| **$T = 151$** |  |  |  |  |  |
| $\mathcal{M}_1$ | -0.519 | 100.000 | 29.571 | 0.222 | 4.803 |
| $\mathcal{M}_2$ | -0.024 | 55.459 | 6.929 | 0.233 | 49.345 |
| $\mathcal{M}_3$ | -0.083 | 66.265 | 8.673 | 0.778 | 96.386 |
| **$T = 165$** |  |  |  |  |  |
| $\mathcal{M}_1$ | -0.298 | 99.123 | 27.636 | 0.122 | 7.895 |
| $\mathcal{M}_2$ | 0.172 | 7.895 | 15.924 | 0.131 | 26.754 |
| $\mathcal{M}_3$ | 0.194 | 11.245 | 19.664 | 0.560 | 69.880 |
| **$T = 190$** |  |  |  |  |  |
| $\mathcal{M}_1$ | -0.238 | 100.000 | 32.210 | 0.156 | 9.705 |
| $\mathcal{M}_2$ | 0.005 | 47.679 | 6.173 | 0.169 | 75.949 |
| $\mathcal{M}_3$ | -0.006 | 52.610 | 6.966 | 0.331 | 98.795 |

#### A3. Results for real data application

**Table 5:** Results of *ℛ*_*t*_ estimates using COVID-19 data in Belgium for the year 2022 with different nowcast dates. The labels *week* and *no week* represent scenarios with and without day-of-the-week effects, respectively.

| | | $\widehat{\mathcal{R}}_t$ | 95% CI lower | 95% CI upper | CI width |
| --- | --- | --- | --- | --- | --- |
| Nowcast Date: March 31, 2022 | $\mathcal{M}_1$ | 0.545 | 0.301 | 0.985 | 0.684 |
| | $\mathcal{M}_2$ - week | 0.965 | 0.702 | 1.326 | 0.624 |
| | $\mathcal{M}_2$ - no week | 0.976 | 0.712 | 1.337 | 0.625 |
| | $\mathcal{M}_3$ - week | 0.926 | 0.528 | 1.625 | 1.097 |
| | $\mathcal{M}_3$ - no week | 0.970 | 0.508 | 1.853 | 1.344 |
| Nowcast Date: April 30, 2022 | $\mathcal{M}_1$ | 0.681 | 0.473 | 0.979 | 0.505 |
| | $\mathcal{M}_2$ - week | 0.836 | 0.647 | 1.079 | 0.432 |
| | $\mathcal{M}_2$ - no week | 0.905 | 0.710 | 1.155 | 0.445 |
| | $\mathcal{M}_3$ - week | 0.849 | 0.569 | 1.266 | 0.697 |
| | $\mathcal{M}_3$ - no week | 0.880 | 0.572 | 1.354 | 0.782 |
| Nowcast Date: May 31, 2022 | $\mathcal{M}_1$ | 0.530 | 0.383 | 0.733 | 0.350 |
| | $\mathcal{M}_2$ - week | 0.855 | 0.705 | 1.037 | 0.332 |
| | $\mathcal{M}_2$ - no week | 0.763 | 0.622 | 0.935 | 0.313 |
| | $\mathcal{M}_3$ - week | 0.838 | 0.614 | 1.145 | 0.531 |
| | $\mathcal{M}_3$ - no week | 0.756 | 0.528 | 1.081 | 0.553 |
| Nowcast Date: June 30, 2022 | $\mathcal{M}_1$ | 0.905 | 0.709 | 1.156 | 0.447 |
| | $\mathcal{M}_2$ - week | 1.104 | 0.927 | 1.314 | 0.387 |
| | $\mathcal{M}_2$ - no week | 1.109 | 0.932 | 1.319 | 0.387 |
| | $\mathcal{M}_3$ - week | 1.078 | 0.849 | 1.369 | 0.521 |
| | $\mathcal{M}_3$ - no week | 1.095 | 0.851 | 1.409 | 0.557 |
| Nowcast Date: July 31, 2022 | $\mathcal{M}_1$ | 0.780 | 0.634 | 0.960 | 0.326 |
| | $\mathcal{M}_2$ - week | 0.899 | 0.747 | 1.081 | 0.335 |
| | $\mathcal{M}_2$ - no week | 0.931 | 0.776 | 1.116 | 0.340 |
| | $\mathcal{M}_3$ - week | 0.908 | 0.731 | 1.128 | 0.398 |
| | $\mathcal{M}_3$ - no week | 0.912 | 0.720 | 1.155 | 0.435 |
| Nowcast Date: August 31, 2022 | $\mathcal{M}_1$ | 0.808 | 0.671 | 0.974 | 0.303 |
| | $\mathcal{M}_2$ - week | 0.943 | 0.812 | 1.095 | 0.283 |
| | $\mathcal{M}_2$ - no week | 0.936 | 0.806 | 1.088 | 0.283 |
| | $\mathcal{M}_3$ - week | 0.914 | 0.749 | 1.116 | 0.367 |
| | $\mathcal{M}_3$ - no week | 0.923 | 0.754 | 1.129 | 0.374 |

**Figure 2:**
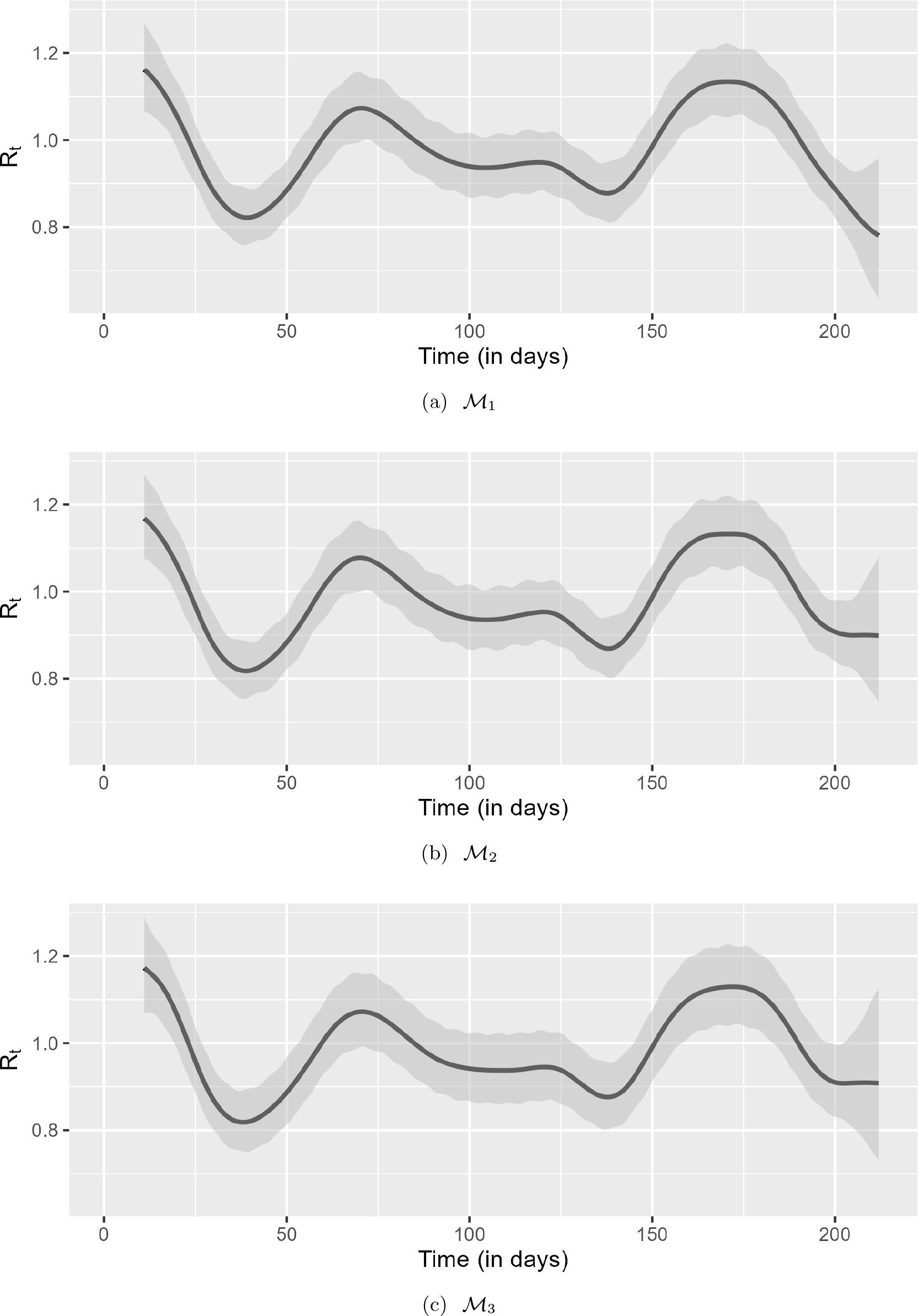
Plot of *ℛ*_*t*_ estimates along with their credible intervals using the COVID-19 data with day-of-the-week effects and nowcast date of July 31, 2022. (a) *ℳ*_1_ - using only reported cases with *estimR()*; (b) *ℳ*_2_ - uses nowcasted incidence with *estimR()*; and (c) *ℳ*_3_ - our approach that jointly models delay and *ℛ*_*t*_.

**Figure 3:**
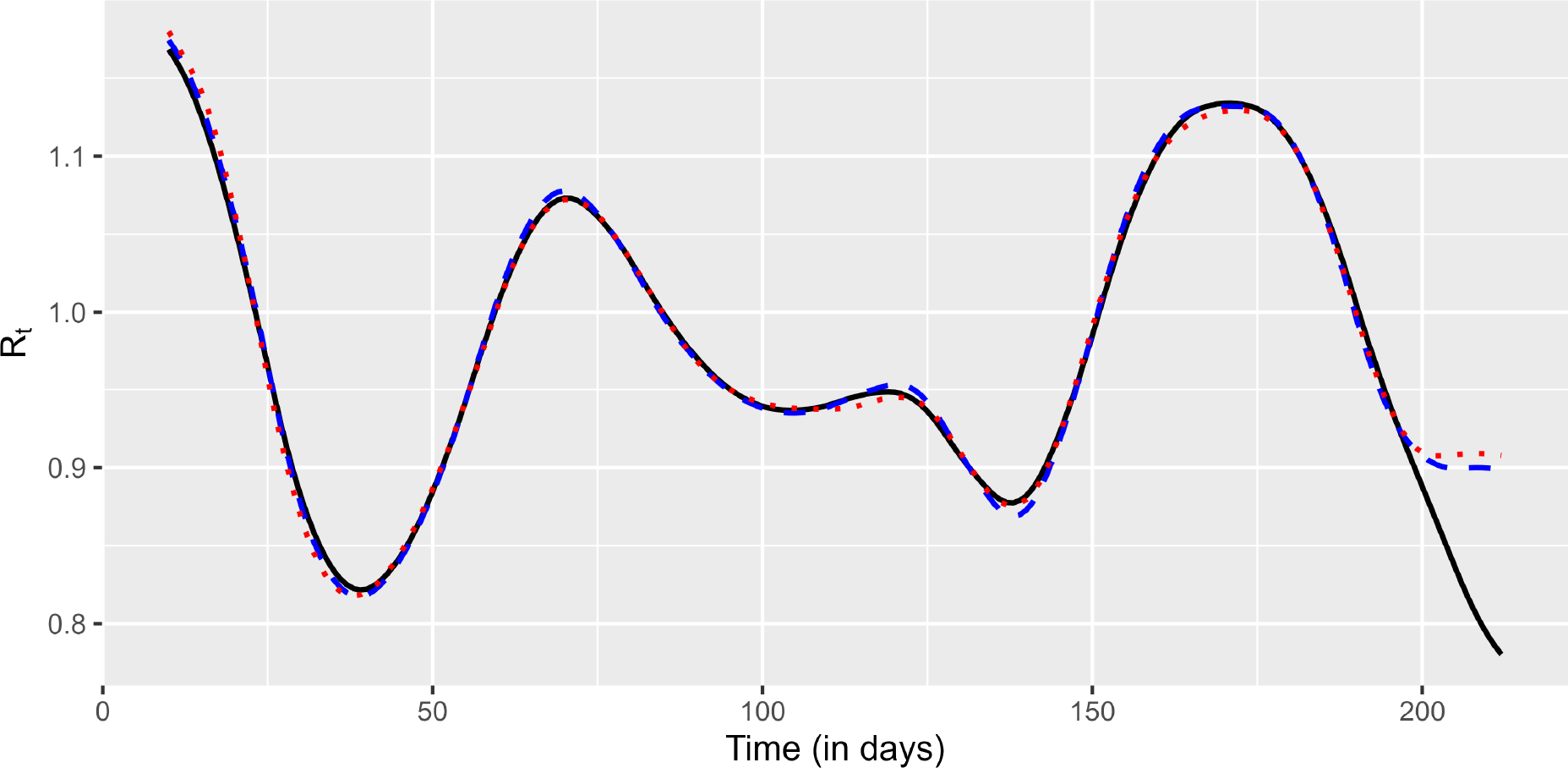
Plot of *ℛ*_*t*_ estimates using the COVID-19 data with day-of-the-week effects and nowcast date of July 31, 2022. *ℳ*_1_ (black line) - uses only reported cases with *estimR()*; *ℳ*_2_ (blue dashed line) - uses nowcasted incidence with *estimR()*; and *ℳ*_3_ (red dotted line) - our approach that jointly models delay and *ℛ*_*t*_.

## References

Abbott, S., Hellewell, J., Thompson, R. N., Sherratt, K., Gibbs, H. P., Bosse, N. I., Munday, J. D., Meakin, S., Doughty, E. L., Chun, J. Y., et al. (2020). Estimating the time-varying reproduction number of SARS-CoV-2 using national and subnational case counts. Wellcome Open Research, 5(112):112.

Eilers, P. H. C. and Marx, B. D. (1996). Flexible smoothing with B-splines and penalties. Statistical Science, 11(2):89–121.

Fraser, C. (2007). Estimating individual and household reproduction numbers in an emerging epidemic. PloS one, 2(8):e758.

Gostic, K. M., McGough, L., Baskerville, E. B., Abbott, S., Joshi, K., Tedijanto, C., Kahn, R., Niehus, R., Hay, J. A., De Salazar, P. M., et al. (2020). Practical considerations for measuring the effective reproductive number, Rt. PLoS Computational Biology, 16(12):e1008409.

Gressani, O. (2021). EpiLPS: a fast and flexible Bayesian tool for estimating epidemiological parameters. [Computer Software]. https://epilps.com/.

Gressani, O., Wallinga, J., Althaus, C. L., Hens, N., and Faes, C. (2022). EpiLPS: A fast and flexible Bayesian tool for estimation of the time-varying reproduction number. PLoS Computational Biology, 18(10):e1010618.

Höhle, M. and an der Heiden, M. (2014). Bayesian nowcasting during the STEC O104: H4 outbreak in Germany, 2011. Biometrics, 70(4):993–1002.

Lang, S. and Brezger, A. (2004). Bayesian P-splines. Journal of Computational and Graphical Statistics, 13(1):183–212.

McGough, S. F., Johansson, M. A., Lipsitch, M., and Menzies, N. A. (2020). Nowcasting by Bayesian smoothing: A flexible, generalizable model for real-time epidemic tracking. PLoS Computational Biology, 16(4):e1007735.

Sumalinab, B., Gressani, O., Hens, N., and Faes, C. (2022). Bayesian nowcasting with Laplacian-P-splines. MedRxiv. 10.1101/2022.08.26.22279249.

Van de Kassteele, J., Eilers, P. H. C., and Wallinga, J. (2019). Nowcasting the number of new symptomatic cases during infectious disease outbreaks using constrained P-spline smoothing. Epidemiology, 30(5):737.

